# Effects of Uninterrupted and Interrupted Sitting on Central Blood Pressure in Chronic Stroke Patients: A Randomized Crossover Trial

**DOI:** 10.64898/2026.09.02.26362083

**Authors:** J. Faulkner, D. Lambrick, S. Fryer, K. Stone, S. Hannah, N. Husted, L. Stoner, A. Mitchelmore

## Abstract

**BACKGROUND:** Sedentary behaviour independently contributes to cardiovascular disease risk. In healthy adults, prolonged uninterrupted sitting acutely elevates blood pressure, whereas light-activity breaks attenuate this response. Whether such strategies confer similar benefits in high-risk populations, such as chronic stroke survivors, remains unanswered.

**OBJECTIVES:** To determine whether non-ambulatory, seated heel-raise interruptions every 10 minutes would attenuate central systolic blood pressure (cSBP) during 3 hours of prolonged sitting in individuals with chronic stroke. We hypothesized that the experimental (heel-raise interruption) condition (EXP) would significantly mitigate the cSBP response relative to uninterrupted sitting (CON).

**METHODS:** Using a randomized crossover design, 15 chronic stroke patients (69.3 ± 10.8 y; 10 male) completed two 3-hour conditions: CON and EXP (10 seated bi-lateral heel raises at 1 rep·s_⁻_¹ every 10 minutes). Central and peripheral blood pressures were assessed pre- and post-each condition using the SphygmoCor XCEL. Condition × Time interactions were tested using repeated-measures ANOVA.

**RESULTS:** The primary hypothesis was confirmed: a significant Condition × Time interaction was detected for cSBP (p < 0.05; _η_p² = 0.28), with EXP eliciting a smaller increase than CON (5.6 mmHg [−6.4, 17.6] vs. 9.6 mmHg [−2.4, 21.6]).

**CONCLUSIONS:** Heel-raise interruptions every 10 minutes attenuate the cSBP response to prolonged sitting in chronic stroke patients and reduce cardiac workload. Frequent, non-ambulatory activity breaks represent a practical secondary prevention strategy for this population, though higher intensity or frequency may be required to fully protect vascular function.

## INTRODUCTION

Sedentary behaviour, defined as time spent in seated, reclined, or lying postures with minimal energy expenditure (≤1.5 METs) (1), has emerged as an independent risk factor for cardiovascular disease (CVD) (2). Prolonged, uninterrupted sitting acutely elevates peripheral systolic blood pressure (pSBP), with meta-analytic evidence demonstrating average increases of approximately 3.2 mmHg, whereas incorporating light physical activity breaks reduces pSBP by around 4.4 mmHg (3). As most research has focused on young, healthy adults (4), the adverse effects of prolonged sitting may be more pronounced in populations at elevated cardiovascular risk, whereby underlying impairments in vascular structure and function could exacerbate the detrimental effects of prolonged sedentary behaviour.

Stroke survivors represent one such high-risk group. As a consequence of their underlying vascular dysfunction, they face substantial risk of recurrent cardiovascular events and secondary stroke (5). Identifying simple strategies to mitigate this risk is particularly important given that chronic survivors spend approximately 75-80% of their waking hours sedentary, with much of this time accumulated in prolonged, uninterrupted bouts (6, 7). In a study of older adults with hypertension and type 2 diabetes, Dempsey and colleagues demonstrated that eight hours of uninterrupted sitting produced clinically significant elevations in pSBP, exceeding those observed in younger, healthy cohorts (8, 9). In ambulatory chronic stroke survivors, 3-minute walking bouts every 30 minutes over an 8-hour period reduced pSBP by 3.5–5.0 mmHg regardless of antihypertensive medication status (10)(16). Yet not all stroke survivors are ambulatory, a reality that demands evaluation of non-standing, seated interruption strategies.

Prior work in this area has focused exclusively on peripheral blood pressure. Unlike peripheral brachial pressure, central aortic systolic blood pressure (cSBP) reflects more closely the pressure directly experienced by the heart, brain, and kidneys, organs central to stroke pathophysiology. Central and peripheral pressures can differ by up to 40 mmHg due to arterial pulse wave amplification, which varies with age, arterial stiffness, and medication class (11)(17); peripheral SBP can therefore misclassify haemodynamic load in older, high-risk individuals. Beyond pSBP, cSBP independently predicts cardiovascular events, left ventricular hypertrophy, and all-cause mortality (11). Although plantarflexor weakness is common after stroke, most ambulatory stroke survivors retain some voluntary ankle plantarflexor function, suggesting that simple plantarflexion-based activities, such as intermittent-heel raises, may be feasible in this population (12)

The aim of this study was therefore to determine whether simple, non-ambulatory light-activity interruptions (experimental condition; EXP) could attenuate the cSBP response to 3 hours of prolonged sitting in a chronic stroke population. We hypothesised that intermittent, bi-lateral, heel raises every 10 minutes would significantly mitigate increases in cSBP compared to uninterrupted sitting (Control condition; CON).

## METHODS

### ETHICS AND REPORTING GUIDELINES

This study is reported in accordance with the Consolidated Standards of Reporting Trials (CONSORT) guidelines (18). The study received institutional ethical approval, conformed to the ethical guidelines of the Declaration of Helsinki, and was registered on the Clinical Trials Registry (NCT03423433). All participants provided written informed consent prior to enrolment.

### PARTICIPANTS

#### RECRUITMENT AND SETTING

Chronic stroke patients were recruited through stroke support groups in the South-Central region of the UK. Participants were eligible if they had a confirmed diagnosis of chronic stroke (>6 months and <10 years post-event), and retained the ability to voluntarily plantarflex. Exclusion criteria included: acute illness at the time of assessment, current smoking, caffeine consumption within 12 hours of testing, and engagement in moderate-to-vigorous physical activity within 48 hours prior to assessment.

### EXPERIMENTAL DESIGN

This was a randomised, counterbalanced crossover trial involving three laboratory visits over a two-week period. Visit 1 served as a familiarisation session; Visits 2 and 3 were EXP and CON, randomly ordered. Each experimental session began between 07:30 and 09:30 am following a minimum three-hour fast (water permitted throughout), with identical individual start times maintained across visits to control for diurnal variation. Ambient temperature and environmental conditions were standardised throughout.

Prior to each visit, participants were reminded of pre-assessment standardisation requirements including fasting for a minimum of three hours, caffeine abstinence for at least 12 hours, and moderate-to-vigorous physical activity restrictions for at least 48 hours before each assessment. Participants were permitted to take morning medication with a small meal to support adherence to the testing schedule. Participants were encouraged to record and repeat the meal prior to subsequent experimental visits. At the start of each visit, participants emptied their bladder and bowel. Participants remained in a seated position, in a comfortable armchair which was used for all assessments, for 3 hours, with measurements recorded following 10 min (BASE) and 3-hours (180 min; POST). Instrumentation was applied to the participants prior to BASE.

#### FAMILIARIZATION

At Visit 1, following written informed consent, participants completed a health history questionnaire and had anthropometric data (height, weight; SECA) recorded. They were then familiarized with all experimental procedures and equipment, including the SphygmoCor XCEL device, blood pressure cuff placement, and the heel-raise protocol with metronome pacing. Cuff placement was maintained on the symptomatic limb across visits for each participant.

### EXPERIMENTAL CONDITIONS

#### EXP (Heel-Raise Interruption)

Participants performed 10 seated bi-lateral heel raises at a cadence of 1 repetition per second, every 10 minutes throughout the 3-hour sitting period. A metronome controlled cadence, and participants were instructed to raise their heels “as high as possible without taking toes off the ground and without discomfort.” The protocol was designed for home use whereby EXP could be embedded during everyday activities such as reading or watching television.

#### CON (Uninterrupted Sitting)

Participants remained seated without any prescribed physical activity interruptions for the full 3-hour period.

#### RANDOMIZATION

We randomized participants to EXP or CON first using a computer-based randomiser (www.randomizer.org). The allocation sequence was generated independently, and condition order was confirmed on the morning of Visit 2. No blinding of participants or research staff was employed, as the nature of the intervention precluded masking.

### EXPERIMENTAL MEASURES

The primary outcome was central systolic blood pressure (cSBP). Descriptive haemodynamic outcomes included peripheral systolic and diastolic blood pressure (pSBP, pDBP), augmentation index (AIx), heart rate (HR), double product (DP), and carotid-femoral pulse wave velocity (cfPWV).

#### PRIMARY OUTCOME — CENTRAL SYSTOLIC BLOOD PRESSURE

cSBP was measured using the SphygmoCor XCEL (AtCor Medical, Sydney, Australia), an oscillometric device with validated transfer function for deriving central aortic waveforms (21). The assessor placed the cuff on the symptomatic limb. Two measures of pulse wave analysis (PWA) were taken at each timepoint; if SBP differed by >5 mmHg or AIx by >4% between readings, a third measurement was recorded and the closest two averaged. Mean arterial pressure (MAP) and heart rate (HR) were simultaneously recorded.

#### DESCRIPTIVE OUTCOMES — HAEMODYNAMIC

The SphygmoCor XCEL additionally measured peripheral blood pressures, heart rate, and central hemodynamics, including blood pressures, and cfPWV. cfPWV was recorded using proximal (carotid) and distal (femoral) pulse pressure waveforms, simultaneously captured using tonometry and an oscillometric cuff, respectively. PWV was calculated by dividing the arterial path length by the pulse transit time (PTT). Path length was determined using a manually constructed caliper device measuring linear distances between the suprasternal notch, carotid artery, and femoral cuff centre line, following recommended manufacturer guidelines (13). cfPWV is recognised as the gold-standard measure of aortic stiffness due to its accuracy, simplicity, and predictive value for cardiovascular events.

### QUALITY CONTROL

#### MEASUREMENT QUALITY CONTROL

All cardiovascular assessments were conducted by a single trained observer to eliminate inter-rater variability. A duplicate measurement protocol with a third measure triggered by pre-specified discordance thresholds (SBP >5 mmHg; AIx >4%) ensured data quality. Averaged values from the two closest readings were used in all analyses.

### STATISTICAL CONSIDERATIONS

#### SAMPLE SIZE

Power analysis was based on sitting-induced changes in cSBP as the primary outcome. Using the pooled standardized mean difference (SMD) from prior meta-analysis of prolonged sitting studies as the effect size of interest (SMD = 0.40) (3), with an alpha of 0.05, power of 0.80, and an assumed within-participant correlation of 0.83 (23), a minimum of 14 participants were estimated to be required. Fifteen participants were enrolled to provide a small buffer against attrition.

#### DATA MANAGEMENT

All raw cardiovascular data were captured and stored within the SphygmoCor XCEL software before export. Data were entered into SPSS v.22 (SPSS, Inc., Chicago, Illinois, USA) for cleaning and analysis. No participants were excluded due to data quality issues, and no missing data were present. No transformations were applied as all variables passed normality testing.

### STATISTICAL METHODS

Statistical analyses were performed using SPSS v.22. Normality was assessed for all variables; all data were normally distributed. Independent sample t-tests were used to assess between-condition differences in baseline values. As no significant baseline differences were detected, repeated-measures ANOVA with two within-participant factors, Condition (EXP vs. CON) and Time (pre [10 min, BASE] vs. post [180min]), were used to test for Condition × Time interactions for all PWA and cfPWV outcomes. A statistical threshold of *P* = 0.10 was used to evaluate Condition × Time interaction effects to account for reduced statistical power (14), whereas a threshold of *P* = 0.05 was used for evaluation of main effects of time or condition.

Data are presented as mean ± standard deviation (SD) and as mean difference between pre- and post-outcomes with 95% confidence intervals. Effect sizes were calculated using partial eta squared (ηp²) with thresholds of 0.01 (small), 0.06 (medium), and 0.14 (large) following Cohen (1969).

## RESULTS

### PARTICIPANTS

All 15 participants completed both conditions and all outcome measure assessments (**Figure 1**), yielding 100% retention and complete data across both the EXP and CON conditions. None withdrew or were excluded after randomization, and no adverse events occurred during either condition.

**Figure 1.**
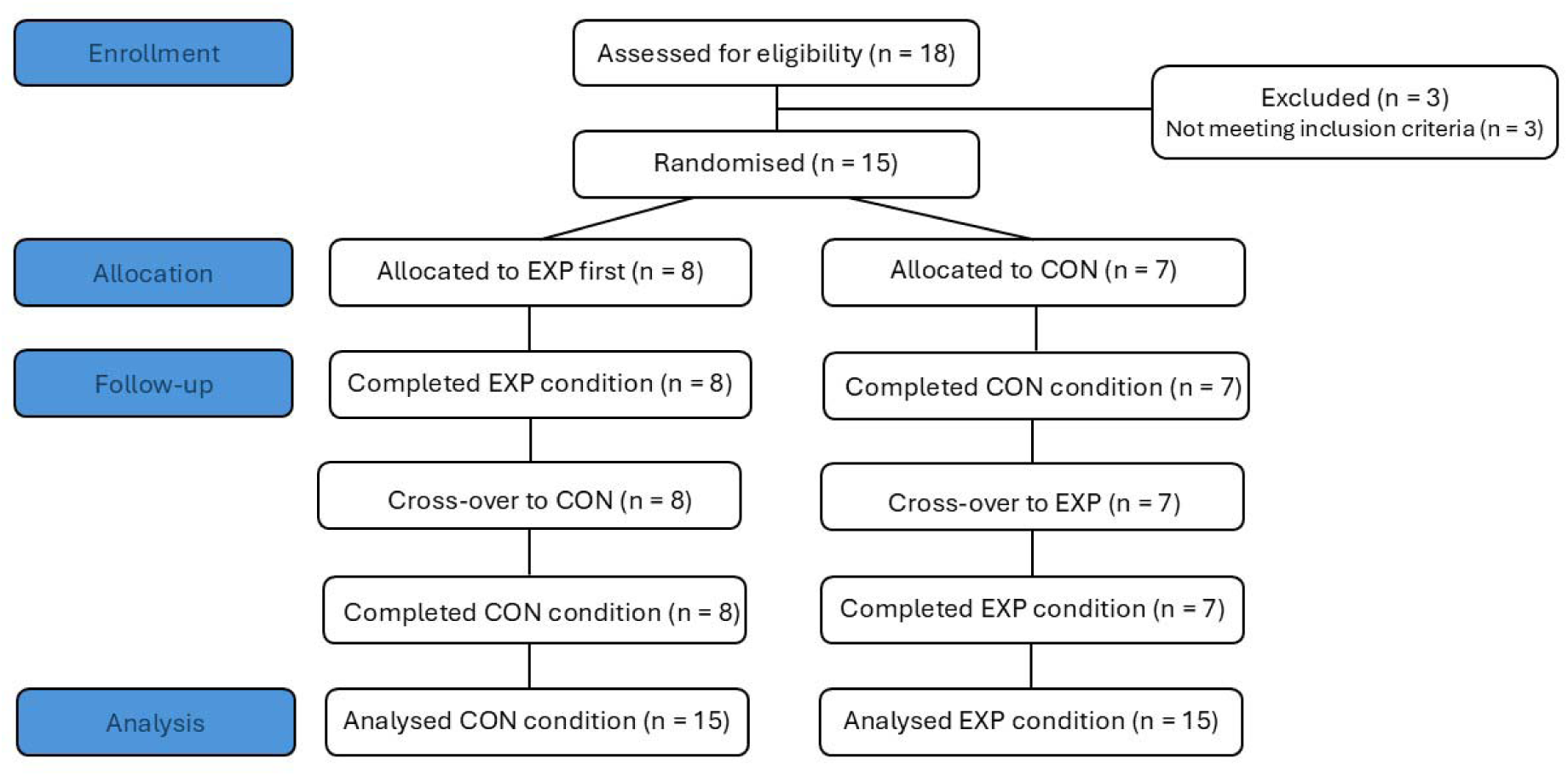
CONSORT Flow Diagram.

### BASELINE CHARACTERISTICS

All participants (69.3 ± 10.8 y, 170.6 ± 8.6 cm, 75.5 ± 9.7 kg) completed the study (**Table 1**). No significant between-condition differences in baseline cardiovascular outcomes were observed (all p > 0.05).

**Table 1.**
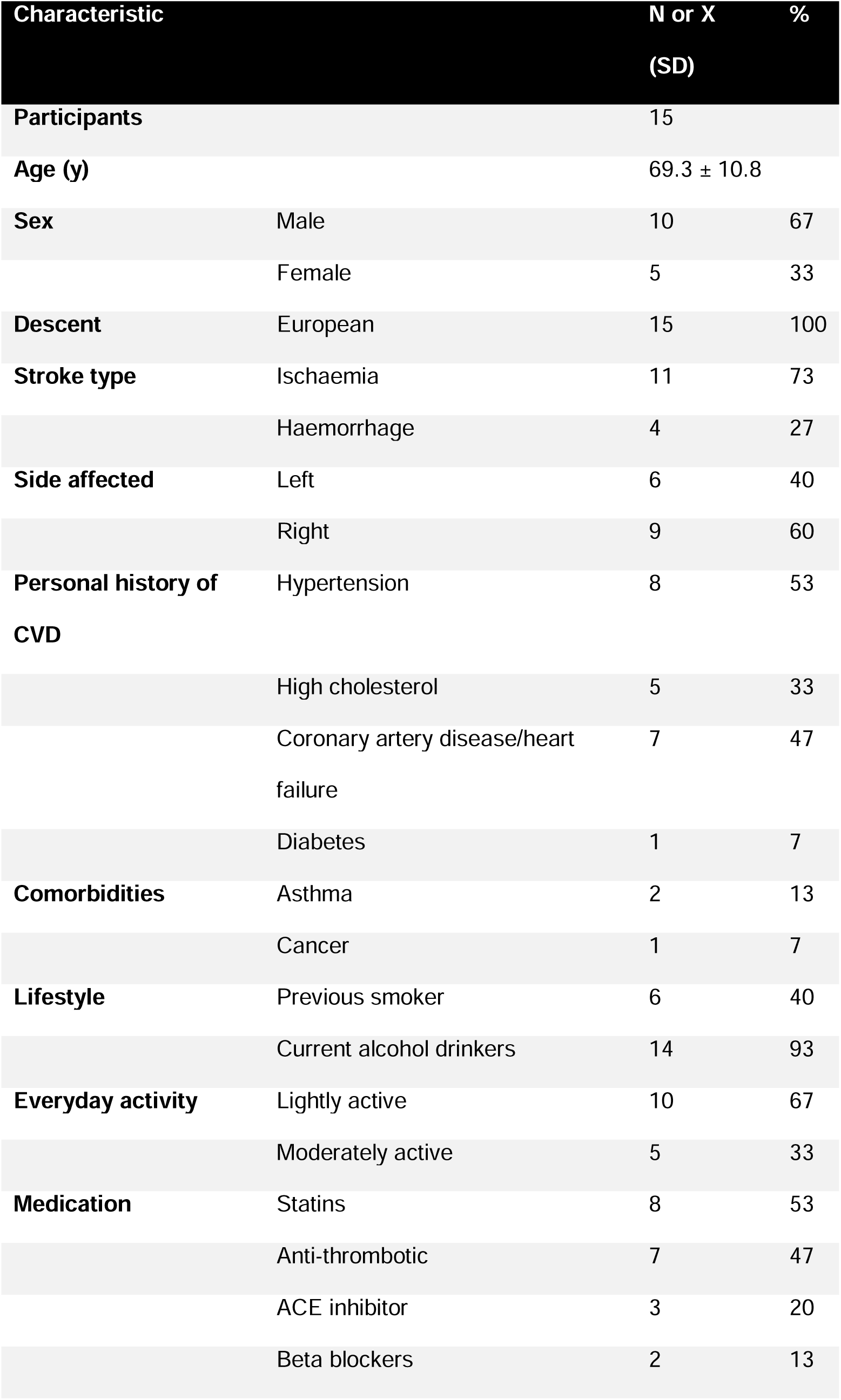
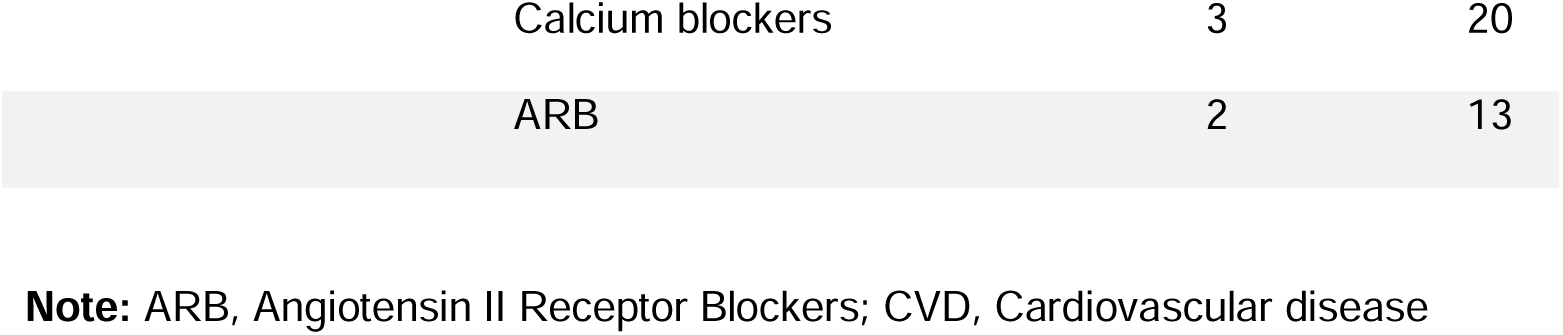
Participant Demographics.

### PRIMARY OUTCOME: CENTRAL SYSTOLIC BLOOD PRESSURE

The primary hypothesis was supported: a significant Condition × Time interaction was detected for cSBP (p < 0.05; ηp² = 0.28; Table 2), with cSBP increasing by 5.6 mmHg [−6.4, 17.6] with EXP versus 9.6 mmHg [−2.4, 21.6] with CON.

**Table 2.** Primary (central systolic blood pressure) and secondary outcomes measures reported for EXP and CON as baseline and post-assessments. Data presented as mean ± SD, including 95% confidence intervals.

|  | Group | BASE | POST | MD (95%CI) | *Condition x Time* |  | *Condition* |  | *Time* |  |
| --- | --- | --- | --- | --- | --- | --- | --- | --- | --- | --- |
| | | | | | *p* | $\eta^2_p$ | *p* | $\eta^2_p$ | *p* | $\eta^2_p$ |
| MAP (mmHg) | IHR | 89 ± 12 | 95 ± 11 | 6.0 (-2.6, 14.6) | 0.43 | 0.06 | 0.99 | 0.01 | **0.01** | 0.49 |
| | CON | 92 ± 15 | 99 ± 14 | 7.0 (-3.9, 17.9) | | | | | | |
| pSBP (mmHg) | IHR | 131 ± 22.7 | 136.2 ± 20.6 | 5.2 (-8.6, 19.0) | **0.02** | 0.32 | 0.48 | 0.04 | **0.02** | 0.312 |
| | CON | 130.2 ± 19.6 | 140.9 ± 22.8 | 10.7 (-2.7, 24.1) | | | | | | |
| pDBP (mmHg) | IHR | 75.9 ± 13.2 | 78.8 ± 9.3 | 2.9 (-5.1, 10.9) | 0.39 | 0.05 | 0.93 | <.01 | **0.05** | 0.26 |
| | CON | 75.1 ± 12.9 | 79.4 ± 11.7 | 4.3 (-4.7, 13.3) | | | | | | |
| cSBP (mmHg) | IHR | 121.8 ± 20.9 | 127.4 ± 19.7 | 5.6 (-6.4, 17.6) | **0.04** | 0.28 | 0.57 | 0.02 | **0.02** | 0.32 |
| | CON | 121.2 ± 18.2 | 130.8 ± 21.4 | 9.6 (-2.4, 21.6) | | | | | | |
| cDBP (mmHg) | IHR | 77.1 ± 13.0 | 79.8 ± 9.3 | 2.7 (-4.9, 10.3) | 0.29 | 0.08 | 0.88 | 0.02 | 0.06 | 0.23 |
| | CON | 76 ± 13.1 | 80.5 ± 11.7 | 4.5 (-4.2, 13.2) | | | | | | |
| cPP | IHR | 44.6 ± 12.5 | 47.6 ± 16.2 | 3.0 (-6.3, 12.3) | 0.21 | 0.11 | 0.47 | 0.04 | 0.14 | 0.15 |
|  | CON | 45.2 ± 13.9 | 50.4 ± 19 | 5.2 (-4.3, 14.7) |  |  |  |  |  |  |
| **Alx (%)** | IHR | 38.0 ± 15.3 | 34.6 ± 13.2 | -3.4 (-13.8, 7.0) | 0.35 | 0.06 | 0.79 | 0.01 | 0.24 | 0.1 |
|  | CON | 36.6 ± 15.1 | 35.4 ± 14 | -1.2 (-11.2, 8.8) |  |  |  |  |  |  |
| **Alx@75 (%)** | IHR | 31.0 ± 16.6 | 25.2 ± 14.6 | -5.8 (-17.2, 5.6) | 0.18 | 0.13 | 0.64 | 0.02 | **0.04** | 0.27 |
|  | CON | 28.4 ± 16 | 26.6 ± 14.7 | -1.8 (-11.9, 8.3) |  |  |  |  |  |  |
| **HR (bpm)** | IHR | 59.4 ± 6.9 | 54.1 ± 7.6 | -5.3 (-10.7, 0.1) | **0.02** | 0.41 | 0.74 | 0.1 | 0.11 | 0.22 |
|  | CON | 57.8 ± 7.1 | 57 ± 10 | -0.8 (-5.9, 4.3) |  |  |  |  |  |  |
| **PTT (m/s)** | IHR | 41.0 ± 5.9 | 39.6 ± 4.7 | -1.4 (-5.4, 2.6) | 0.33 | 0.19 | 0.65 | 0.05 | 0.21 | 0.29 |
|  | CON | 44.4 ± 15.1 | 37.3 ± 9 | -7.1 (-17.0, 2.8) |  |  |  |  |  |  |
| **cfPWV (m/s)** | IHR | 11.5 ± 1.9 | 12.0 ± 1.8 | 0.5 (-0.9, 1.9) | 0.34 | 0.18 | 0.94 | <0.01 | 0.19 | 0.32 |
|  | CON | 11.4 ± 2.6 | 12.5 ± 2.8 | 1.1 (-0.8, 3.0) |  |  |  |  |  |  |
| **pDP (HR x SBP)** | IHR | 7398 ± 1146 | 7236 ± 1663 | -162 (-1230, 906) | **0.01** | 0.55 | 0.79 | 0.01 | 0.38 | 0.07 |
|  | CON | 7096 ± 1185 | 7651 ± 1544 | 555 (-474, 1584) |  |  |  |  |  |  |

**Note:** Alx, Augmentation index; Alx75, Alx @ 75 beats per minute; cfPWV, Carotid-femoral pulse wave velocity; cDBP, Central diastolic blood pressure; CI, Confidence interval; cSBP, Central systolic blood pressure; DBP, Peripheral diastolic blood pressure; pDP, Peripheral Double product; HR, Heart rate; MAP, mean arterial pressure; PTT, Pulse transit time; pSBP, Peripheral systolic and diastolic blood pressure

### DESCRIPTIVE OUTCOMES: HAEMODYANMIC

The secondary outcomes contextualised and reinforced the primary cSBP result. Significant Condition × Time interactions were detected for pSBP, HR, and pDP (all p < 0.05; ηp² = 0.32–0.55; **Table 2**). The pSBP response mirrored the primary outcome, with EXP eliciting a mitigated increase (MD [95%CI]: 5.2 mmHg [−8.6, 19.0]) compared with CON (10.7 mmHg [−2.7, 24.1]). HR and pDP responses further support a reduced myocardial oxygen demand with EXP: HR decreased more with EXP (−5.3 bpm [−10.7, −0.1]) than CON (−0.8 bpm [−5.9, 4.3]), and pDP decreased with EXP (−162 [−1230, 906]) while CON produced an increase (555 [−474, 1584]). Time main effects for pDBP and MAP (both p < 0.05; ηp² = 0.26–0.49) indicate sitting-induced elevations irrespective of condition.

## DISCUSSION

### SUMMARY OF KEY FINDINGS

This study assessed whether a simple, non-ambulatory light-activity interruptions (specifically intermittent seated heel raises) could attenuate the adverse cSBP response to 3 hours of prolonged sitting in individuals with chronic stroke. Our study demonstrated that frequent heel-raise interruptions every 10 minutes significantly lowered the sitting-induced increases in cSBP **(Figure 2).**

**Figure 2.**
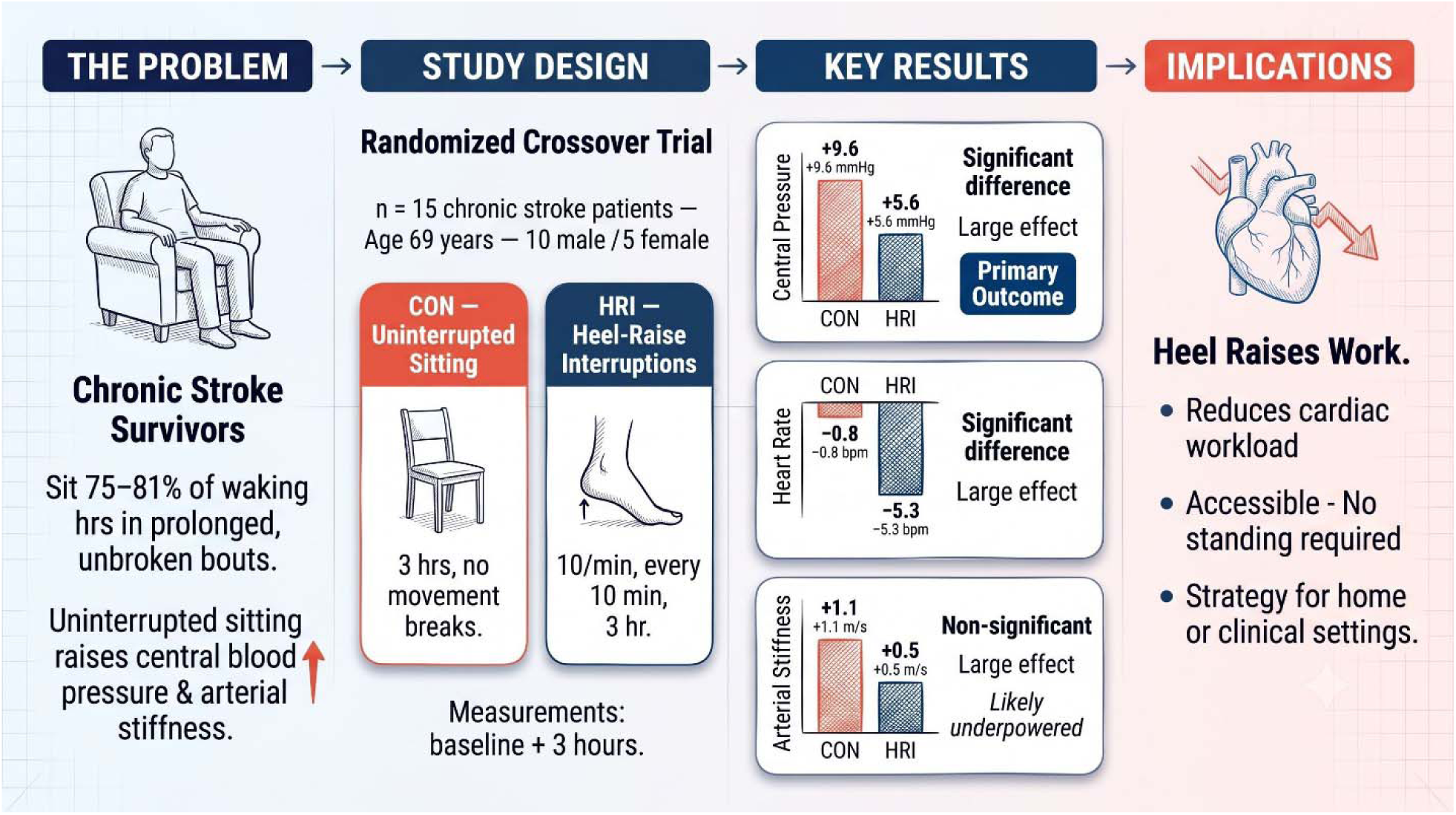
Graphical abstract.

### LIMITATIONS AND STRENGTHS

This study has limitations, though each reflects a deliberate design choice appropriate to its pilot nature. The absence of a non-stroke comparison group limits causal inference regarding stroke-specific mechanisms; however, the study was intended to assess the intervention’s efficacy within a stroke population rather than against a non-stroke control group. The sample was exclusively European, older, and predominantly male, a known constraint of single-center, pilot recruitment that limits generalizability. A single pre-post design precludes characterization of the haemodynamic trajectory across the sitting period. Laboratory conditions maximize internal validity but may not capture real-world variability. Physical fitness and habitual activity were not quantified, though prior evidence suggests these do not materially affect sitting-induced haemodynamic responses (38). The small sample limits statistical power for secondary outcomes, particularly cfPWV, and precludes subgroup analyses.

Despite the above, the design carries notable strengths. A randomized crossover design with tightly controlled experimental conditions (standardized posture, fasting, session timing, and ambient environment) maximized internal validity. Complete data across all 15 participants (100% retention with no missing data points) supports the reliability of all findings. The may be directly applicable to home-based secondary prevention, strengthening the translational relevance and potential adherence.

### COMPARISON TO LITERATURE

The finding that cSBP elevations during prolonged sitting are attenuated by light-activity interruptions is consistent with, and extends, evidence from clinical populations. Fryer and colleagues demonstrated that sit-to-stand, calf raises, and walking interruptions reduced a 15 mmHg sitting-induced pSBP rise to 4 mmHg in coronary heart disease patients (15), the closest prior analogue to the present study. Our findings replicate this pattern in stroke survivors, with CON producing 10–11 mmHg increases in cSBP versus approximately half that magnitude with EXP. This aligns with meta-analytic evidence that the largest pSBP elevations occur within the first few hours of sitting (4). Although the intervention attenuated blood pressure responses, it did not change arterial stiffness (cfPWV), suggesting that the stimulus provided by intermittent heel raises was insufficient to counter non-blood pressure-mediated mechanisms contributing to vascular stiffening, such as endothelial dysfunction. Furthermore, increases of >5 mmHg in both cSBP and pSBP were still observed in the EXP condition, indicating that the activity, and/or the volume performed, was insufficient to fully offset the adverse haemodynamic effects of prolonged sitting.

Three contributions extend the existing literature. First, this is the first sitting interruption study in stroke patients which has shown significant changes in central blood pressure. Second, prior sitting-interruption studies have typically required ambulatory capacity (e.g., standing or walking), whereas this work demonstrates some haemodynamic protection using a wholly seated strategy, extending the evidence base to stroke survivors regardless of mobility status. Importantly, such a seated approach may also be more behaviourally acceptable and feasible for individuals with mobility limitations, potentially supporting greater adherence in real-world settings. Third, participants here represent a higher-risk group than those enrolled in prior sitting interruption trials, which have predominantly included healthy or pre-clinical cohorts (3, 4). That a simple, low-intensity intervention attenuates cSBP in this population strengthens the translational case for non-ambulatory activity breaks as a secondary prevention tool. The HR and pDP responses indicate reduced myocardial oxygen demand with EXP, further supporting the cardioprotective model proposed by Stoner *et al* (16).

cfPWV did not show a significant Condition × Time interaction, though the effect size was large (ηp² = 0.18) and the CON-condition increase of 1.1 m/s exceeded the clinically significant threshold (17). This pattern likely reflects impaired vasodilatory capacity in this post-stroke sample. Endothelial dysfunction blunts the flow-mediated responses to light movement that underpin cfPWV attenuation in healthy cohorts (18–20). These data were collected to contextualize the cSBP response. Future studies will need adequate power to determine whether higher-intensity or more frequent interruptions can fully protect aortic stiffness in this population.

### IMPLICATIONS

Prior to this study, evidence for sitting interruption strategies in stroke was confined to ambulatory walking-based approaches assessed over extended sitting periods. This study demonstrates that a simple, non-ambulatory seated activity (intermittent heel raises) provides meaningful haemodynamic protection during a clinically relevant 3-hour sitting bout, without requiring standing capacity, exercise equipment, or supervision. For the large proportion of stroke survivors who are lightly active or face barriers to physical activity, this strategy offers a practical approach to cardiovascular secondary prevention at home or in community settings (6, 7, 21). The attenuation of cSBP by approximately 5–6 mmHg with EXP versus CON approaches the 5 mmHg threshold considered clinically significant for cardiovascular risk reduction (6), supporting the translational relevance of this approach. Future research should explore dose-response relationships (i.e., varying the frequency, duration, and intensity of interruptions) to determine the optimal prescription for preserving both haemodynamic and vascular function. Longer-term trials are needed to examine repeated daily interruption strategies and mechanistic pathways, including shear stress and endothelial function (19, 22). Studies enrolling women and participants across varied stroke severity will be essential to establish generalisability and clinical utility.

## CONCLUSIONS

Prolonged uninterrupted sitting for 3 hours adversely affects cardiovascular function in individuals with chronic stroke. Interrupting sitting every 10 minutes with brief, light-intensity seated heel raises attenuates cSBP and pSBP, and reduces cardiac workload as indexed by HR and pDP. Frequent, non-ambulatory activity breaks represent a practical and evidence-supported secondary prevention strategy for stroke survivors, though greater or more intense stimuli may be required to fully protect vascular health in this population.

## Data Availability

All data produced in the present work are contained in the manuscript

## ACKNOWLEDGEMENTS AND ETHICAL STATEMENTS

### Data Availability

Data are available from the corresponding author upon reasonable request.

## Acknowledgments

The authors thank the participants and stroke support groups for their involvement. We would also like to thank Simon Jobson for co-supervising Andrew Mitchelmore’s PhD.

## Author Contributions (CRediT)

Faulkner, J.: Conceptualization, Methodology, Investigation, Writing Original Draft; Lambrick, D.: Methodology, Investigation; Fryer, S.: Formal Analysis, Writing, Review & Editing; Stone, K.: Formal Analysis, Writing: Review & Editing; Hannah, S.: Investigation, Review & Editing; Hudson, N.: Investigation, Review & Editing; Stoner, L.: Conceptualization, Supervision, Writing, Review & Editing; Mitchelmore, A.: Investigation, Writing, Review & Editing.

## Funding

n/a.

## Ethics Approval

The study received institutional ethical approval, conformed to the Declaration of Helsinki, and was registered on the Clinical Trials Registry (NCT03423433).

## Conflicts of Interest

None declared.

## Notes

### Competing Interest Statement

The authors have declared no competing interest.

### Clinical Trial

NCT03423433

### Author Declarations

Ethics committee at the University of Winchester gave ethical approval for this work

